# Sample Pooling as a Strategy of SARS-COV-2 Nucleic Acid Screening Increases the False-negative Rate

**DOI:** 10.1101/2020.05.18.20106138

**Authors:** Yichuan Gan, Lingyan Du, Oluwasijibomi Damola Faleti, Jing Huang, Gang Xiao, Xiaoming Lyu

**Author notes:** These authors contribute equally to this work (Yichuan Gan, Lingyan Du, Oluwasijibomi Damola Faleti). **Corresponding Author:** Xiaoming Lyu, MD., PhD.(;) or Gang Xiao, MD., PhD. Department of laboratory medicine, The Third Affiliated Hospital, Southern Medical University, No. 183, Zhongshan Avenue West, Guangzhou, China. 510630.

## Abstract

**Background:** Identification of less costly and accurate methods for monitoring novel coronavirus disease 2019 (CoViD-19) transmission has attracted much interest in recent times. Here, we evaluated a pooling method to determine if this could improve screening efficiency and reduce costs while maintaining accuracy in Guangzhou, China.

**Methods:** We evaluated 8097 throat swap samples collected from individuals who came for a health check-up or fever clinic in The Third Affiliated Hospital, Southern Medical University between March 4, 2020 and April 26, 2020. Samples were screened for CoViD-19 infection using the WHO-approved quantitative reverse transcription PCR (RT-qPCR) primers. The positive samples were classified into two groups (high or low) based on viral load in accordance with the CT value of COVID-19 RT-qPCR results. Each positive RNA samples were mixed with COVID-19 negative RNA or ddH2O to form RNA pools.

**Findings:** Samples with high viral load could be detected in pool negative samples (up to 1/1000 dilution fold). In contrast, the detection of RNA sample from positive patients with low viral load in a pool was difficult and not repeatable.

**Interpretation:** Our results show that the COVID-19 viral load significantly influences in pooling efficacy. COVID-19 has distinct viral load profile which depends on the timeline of infection. Thus, application of pooling for infection surveillance may lead to false negatives and hamper infection control efforts.

**Funding:** National Natural Science Foundation of China; Hong Kong Scholars Program, Natural Science Foundation of Guangdong Province; Science and Technology Program of Guangzhou, China.

**Research in context:** *Evidence before this study:* Since it emergence in late 2019, CoViD-19 has dramatically increased the burden healthcare system worldwide. A research letter titled “Sample Pooling as a Strategy to Detect Community Transmission of SARS-CoV-2” which was recently published in JAMA journal proposed that sample pooling could be used for SARS-COV-2 community surveillance. Currently, the need for large-scale testing increases the number of 2019-nCOV nucleic acid analysis required for proper policy-making especially as work and normal school resumes. As far as we know, there are many countries and regions in the world, who are beginning to try this strategy for nucleic acid screening of SARS-CoV-2.

*Added value of this study:* We carried out a study using pooled samples formed from SARS-COV-2 negative samples and positive samples with high or low viral and assessed detection rate for the positive samples. We found that positive sample with high viral load could be detected in pools in a wide range of dilution folds (ranging from1/2 to 1/50). On the contrary, the sample with low viral load could only be detected in RNA “pools” at very low dilution ratio, and the repeatability was unsatisfactory. Our results show the application of the “pooling” strategy for large-scale community surveillance requires careful consideration and depends on the viral load of the positive samples.

*Implications of all the available evidence:* Although the number of newly diagnosed cases has been reducing in some parts of the world, the possibility of a second wave of infection has made quick and efficient data gathering essential for policy-making, isolation and treatment of patients. Fast and efficient nucleic acid detection methods are encouraged, but sample pooling as a strategy of SARS-COV-2 nucleic acid screening increased the false-negative rate, especially those with asymptomatic infections have lower viral load. Therefore, the application of the “pooling” strategy for large-scale community surveillance requires careful consideration by policy makers.

## Introduction

Since it emergence in late 2019, CoViD-19 has dramatically increased the burden healthcare system worldwide. As of April 26, 2020, about 2.8 million diagnosed cases, and 198 thousand confirmed deaths have been reported globally. CoViD-19 is characterized by fever and severe acute respiratory syndrome, which may be accompanied by gastrointestinal and hepatic dysfuntion(1, 2). Although the number of newly diagnosed cases has been reducing in some parts of the world, the possibility of a second wave of infection has made quick and efficient data gathering essential for policy-making, isolation and treatment of patients. (3) The need for large-scale testing increases the number of 2019-nCOV nucleic acid analysis required for proper policy-making especially as work and normal school resumes

Strategies that reduce the cost of tracking infections could enhance proper monitoring, especially in regions faced with limited inspection personnel and equipment. One such strategy is the pooled testing of samples. It has been applied for community surveillance of other microbial infections including chlamydia trachomatis and dichelobacter nodosus. (4, 5) However, it has not been applied for globally for community screening for 2019-nCOV infections. A research team from Stanford University School of Medicine proposed sample “pooling” for tracking of community infection.(6) The authors showed that the sample “pooling” strategy improves the overall detection efficiency and may enhance prevention and control measures.

CoViD-19 infection has two phases (late and early stages); each with a unique viral load profile. To ascertain the efficacy of pooling for tracking 2019-nCOV infections, we build the RNA sample “pooling” strategy which is involving mixing positive RNA sample with low or high viral load with multiple negative RNA samples together.

## Methods

### Sample collection

Patients who came for routine medical examination or fever were recruited for the study at the Third Affiliated Hospital of Southern Medical University, Guangzhou City, China. Throat swabs were collected by doctors in fever clinics and isolation wards. The swab was extended into the patient’s cavity, wiped around the tonsils and the mucosa of the posterior wall till thoroughly infiltrated. Then, the swab was inserted into a test tube which contained 2 ml of the transport swab buffer mixed with lysis buffer. The samples at 2-8□ and delivered to a medical laboratory in 2 hours. Using an RNA extraction kit (Shanghai ZJ Bio-Tech), total RNA was extracted. We obtained swab samples tested between March 4 to April 26, 2020.

### Individual RT-PCR tests in the medical laboratory

RT-qPCR was performed in the medical laboratory to detected the 2019-nCOV nucleic acid RNA with QIAamp Viral RNA Mini Kit (QIAGEN Biotech) in a SLAN-96P qPCR machine (Sansure Biotech) using WHO primers and probe (E_Sarbeco_R: ATATTGCAGCAGTACGCACACA, E_Sarbeco_F: ACAGGTACGTTAATAGTTAATAGCGT, E_Sarbeco_P: ACACTAGCCATCCTTACTGCGCTTCG). The reactions were carried out in 25ul mixtures that contained 19 ul of detection mix, 1ul of RT-qPCR polymerase and 5ul of RNA template. The reaction was heated to 45□ for 10 minutes for reverse transcription, denatured in 95□ for 90 seconds, and then the experimental conditions consisted of the following cycles: amplification was carried in 95□ for 3 seconds and 58□ for 20 seconds (45 cycles).

### RNA samples “pooling.”

Three positive samples; two with high viral load (Case 1 and Case 2) and one with low viral load (Case 3), and 8094 negative samples were used for the study. The positive RNA sample Case1 or Case 2 was mixed with equal volumes of 1, 4, 9, 19, 49, 99, 199, 499, 999 negative RNA samples in order to dilute into 1:2, 1:5, 1:10, 1:20, 1:50, 1:100, 1:200, 1:500, 1:1000. Using double distilled water (ddH_2_O), other dilution ratio of 1:2000, 1:4000, 1:8000 were made. The third positive RNA sample, Case 3, was mixed with equal volumes of 1, 4, 9, 19, 49, 99, 199 negative RNA samples to make 1/2, 1/5, 1/10, 1/20, 1/50, 1/100, 1/200 dilution. Each RNA “pool” was detected using RT-qPCR test as one RNA template.

### Pooled RNA samples for RT-qPCR

RT-qPCR procedure for individual samples was performed using the same reagent/machine and analyzed using the blind method. Dr. Yichuan Gan mixed the positive RNA sample with negative RNA samples to dilute different ratio. *Miss* Lingyan Du was responsible for detecting the RNA “pool” using RT-qPCR with unknown of the dilution ratio. Finally, Dr. Xiaoming Lyu summarised and analyzed the data.

### Role of the funding source

The funding organization(s) played no role in the design and conduct of the study; collection, management, analysis, and interpretation of the data; preparation, review, or approval of the manuscript; and decision to submit the manuscript for publication.

## Results

The patient Case 1 had a fever with body temperature at 39.3 degrees Celsius (L), which was accompanied with headache and muscle soreness, and a throat swab sample was collected timely. Case 2 had abdominal pain for two days with l diarrhoea (three to four times each day). The patient also had low-grade fever of 37.3□, and a throat swab sample was collected after a physical examination. They both showed significant positive results of three CoViD-19 probe genes. The positive RNA sample was diluted into 1:2, 1:5, 1:10, 1:20, 1:50, 1:100, 1:250, 1:500, 1:1000 to make RNA “pools”. We tested RNA “pools” using RT-qPCR, then found that the positive RNA samples could still be observed in pools of up to 1:1000 ratio, even to 1:8000 diluted in ddH2O with Case 1 (Figure 1A and 1B).

**Figure 1.**
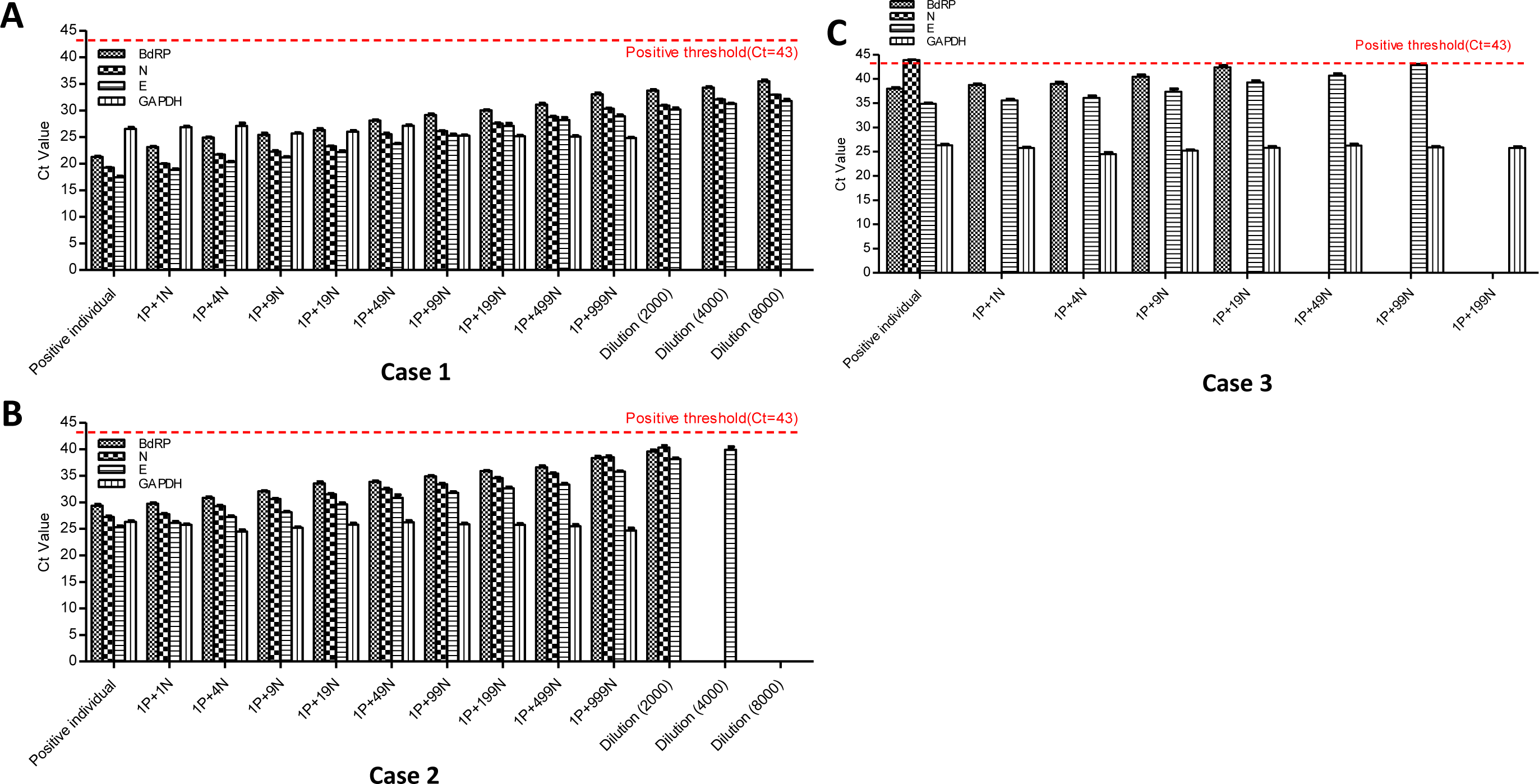
RT-PCR CT values of the three COVID-19 positive samples and their respective pools formed with negative samples. Three positive samples; two with high viral load (Case 1 and Case 2) and one with low viral load (Case 3), and 8094 negative samples were collected from individuals who came for a health check-up or fever clinic in The Third Affiliated Hospital, Southern Medical University between March 4, 2020 and April 26, 2020. The positive RNA sample Case was mixed with equal volumes of 1, 4, 9, 19, 49, 99, 199, 499, 999 negative RNA and 1:2000,1:4000,1:8000 diluted in ddH2O with Case 1 and Case 2.

The patient Case 3 had a fever for a week followed by coughing (with sputum) and sore throat. The body temperature was between 37.1□ and 38□. RT-qPCR analysis showed positive result in only two of CoViD-19 probe genes. This positive RNA sample was diluted into the “pool” of one sample to make a 1/2, 1/5, 1/10, 1/20, 1/50, 1/100 dilution. The mixed RNA “pools” were analyzed using the CoViD-19 primers. The positive sample with low viral load could only be observed at a low dilution ratio (which ration; p<0.5) (Figure 1C).

Each positive RNA sample was diluted with different negative RNA samples (1/2, 1/5, 1/10, 1/20, 1/50). For each dilution ratio, we had ten repeats which was analyzed for the CoViD-19 primer probe. The mixed RNA “pools” formed from the Case 1 and Case 2 samples could be detected each time when the 1/2, 1/5, 1/10, 1/20 RNA pools were used and 90 percent of time when 1/50 ratio RNA pools were used (Figure. 2A). In contrast, the positive detection rate in Case 3 was unsatisfactory. As the dilution ratio increase, the detection became negligible (Figure. 2A).

**Figure 2.**
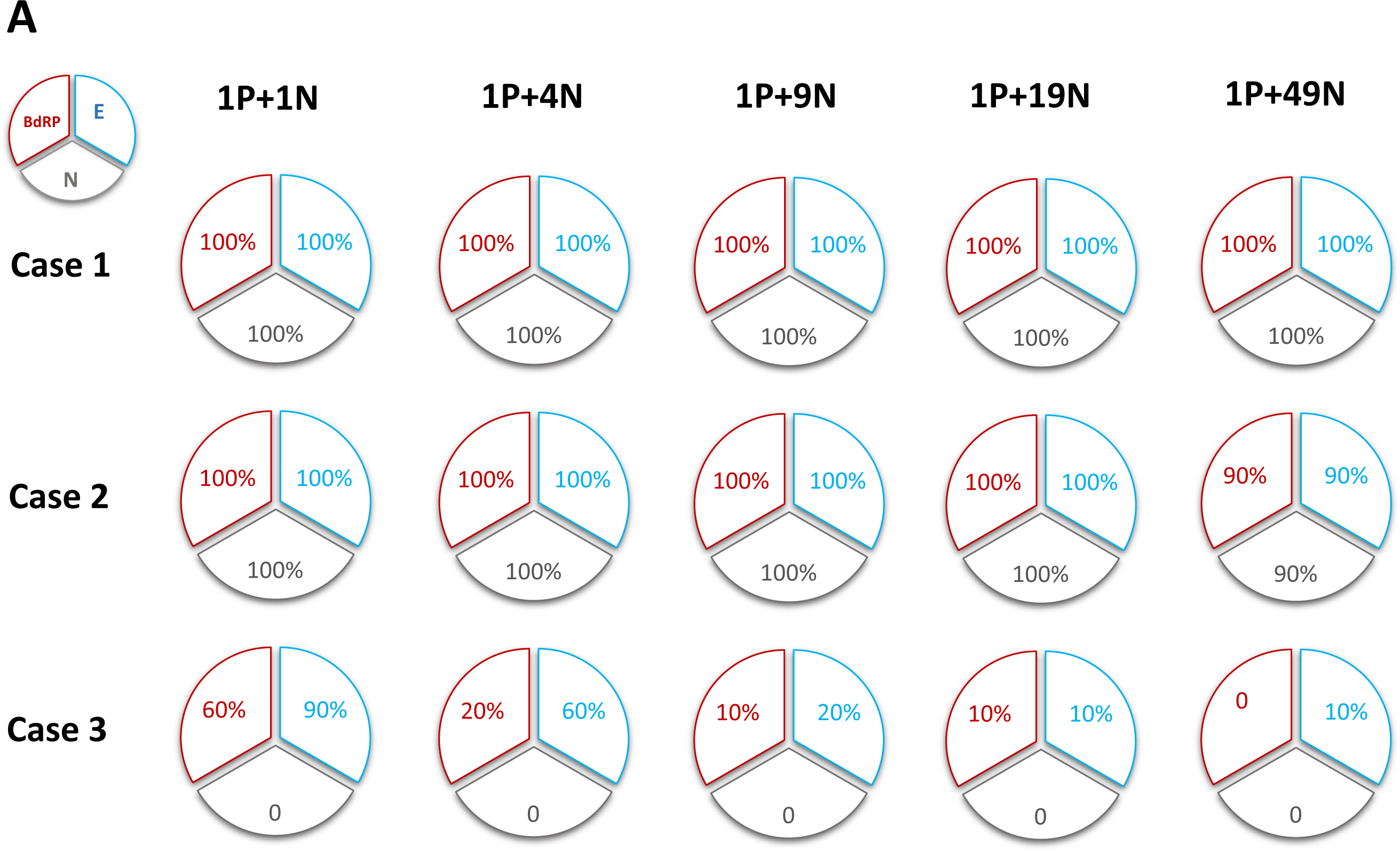
The number of times the positive sample could be detected in a pooled RNA mixture. Each test was carried ten times.

## Discussion

The long-term curtailment of COVID 19 worldwide depends on quick and efficient monitoring of patients, and affordability of testing in resource-constrained countries. Currently available commercial assays are efficient; however, they require specialized equipment, which may be limited in supply in the face of the growing need for mass testing. Pooled testing could be implemented to drastically reduce the overall test costs and reduce waiting time; thereby enhancing infection survelliance. In our study, RNA pooling strategy was used to examine the detection rate of positive COVID 19 samples collected at different phases of infection in a pool of negative samples.

Recent reports have shown that the 2019-nCOV viral load in posterior oropharyngeal saliva samples was highest during the first week of symptom onset and then declined steadily for most patients.(7, 8) We classified the positive samples into two cases; high viral load and low viral load in accordance to the CT value of COVID-19 qPCR results and, the number of days after symptom onset (the time of sample collection). Using negative RNA samples or ddH_2_O, the positive RNA samples were diluted into different ratio to making RNA “pools”. We then carried out COVID-19 qPCR in each reconstituted RNA sample “pools”. Our results showed that samples from positive patients with an earlier symptom onset and a higher viral load, could be in RNA “pool” a wide range of dilution folds (ranging from 1/2 to 1/100 with a positive detection rate of p>0.05)). On the contrary, sample from positive patients with low viral load and late symptom onset could only be detected in RNA “pools” at very low dilution, and the repeatability was unsatisfactory.

A number of recent studies have proposed samples pooling for the tracking COVID-19 spread so as to increase testing rate and enhace policy making. In line with recent research, we show that samples from positive patients with high viral load, can be detected in pool negative sanples. However, detecting samples from positive patients with low viral load in a pool was diificult and may lead false negatives if “pooling” strategy is applied community-wide. Although the shortage of detecting equipment and personnel may make sample pooling a good strategy in resource-strained areas, the high rate of false negatives may undermine efficient tracking and good policy making. Thus, application of the “pooling” strategy for large-scale community screening requires careful consideration.

## Data Availability

All data referred to in the manuscript is available.

## Author Contributions

Dr. Yichuan Gan mixed the positive RNA sample with negative RNA samples to dilute different ratio. *Miss* Lingyan Du was responsible for detecting the RNA “pool” using RT-qPCR with unknown of the dilution ratio. Dr. Xiaoming Lyu and Dr. Oluwasijibomi Damola Faleti summarised and analyzed the data. Dr. Yichuan Gan, *Miss* Lingyan Du and Dr. Oluwasijibomi Damola Faleti contribute equally to this work.

Concept and design: Dr. Xiaoming Lyu.

Acquisition, analysis, or interpretation of data: All authors.

Statistical analysis: Dr. Yichuan Gan and Dr. Oluwasijibomi Damola Faleti.

Drafting of the manuscript: Dr. Yichuan Gan and Dr. Oluwasijibomi Damola Faleti. Administrative, technical, or material support: Dr. Gang Xiao and Mr. Jing Huang. Obtained funding: Dr. Xiaoming Lyu.

## Disclaimer

The findings and conclusions in this report are those of the authors and do not necessarily represent the official position of the Centers for Disease Control and Prevention.

## Declaration of interests

None reported.

## Acknowledgments

This work was funded by grants from National Natural Science Foundation of China (No. 81502335), Hong Kong Scholars Program (XJ2017-146), Natural Science Foundation of Guangdong Province (No.2020A1515010081). Science and Technology Program of Guangzhou, China (No. 201704020127).

## Notes

### Competing Interest Statement

The authors have declared no competing interest.

### Author Declarations

The recruitment process, delivery and use of clinic samples obtained from patients who had fever or routine medical examination, and the experimental procedures were approved by the Medical Ethics Committee of the Third Affiliated Hospital Of Southern Medical University, Guangzhou, China.

